# Aggregation and analysis of 25 years of prion disease natural history extracted from published literature

**DOI:** 10.64898/2026.08.07.26359973

**Authors:** Leo M.H. Xu, Daniel A. Sprague, Sonia M Vallabh, Eric Vallabh Minikel

## Abstract

**Background and Objectives:** Prion disease is an untreatable, typically rapidly progressive dementia. New drug candidates designed to lower prion protein are now entering clinical trials. To support future pivotal trials, we sought to assemble and analyze a public dataset of natural history data characterizing the symptomatic course of prion disease in order to provide a quantitative basis for modelling and rational trial design.

**Methods:** We extracted data from medical publications between 2000 and 2024 reporting on n≥5 prion disease patients. Information about demographics, ascertainment, and clinical milestones, such as time from onset to death, akinetic mutism, diagnostic testing and outcomes, were extracted for aggregate cohorts of ≥2 patients and individual patient-level data (PLD). We computed summary statistics of cohorts and PLD, visualized survival through forest plots and Kaplan-Meier curves, analyzed covariates regarding survival, and identified biases and heterogeneity within the literature.

**Results:** From 245 included publications we extracted 418 aggregate cohort medians and 1,400 rows of individual PLD. 90% of cohorts and 91% of individual patients had symptom-to-death milestones, while only 7% and 11% had a time interval from a clinical presentation milestone to death, respectively. Symptom-to-death intervals varied as much as 3.2-fold even between cohorts of the same histopathologic subtype, and akinetic mutism occurred 73% sooner than death when both endpoints were reported (N=53). Indicators of disease severity, such as a cognitive test, were rarely present in either aggregate data (11%) or PLD (6%).

**Discussion:** Data routinely reported in publications can quantify diagnostic delay and covariates affecting survival time, but are limited in ability to inform pivotal trial design because most such data are aggregated, cross-sectional, lack indicators of disease severity, and present timelines beginning with onset rather than more relevant clinical milestones, such as diagnosis, that may better reflect the moment of potential for trial enrollment. There is a need for clinical data to report milestones such as intervals from diagnosis to death, for longitudinal cognitive and functional scores, and for deposition of publicly accessible PLD.

## Introduction

Natural history data play an important role in the development of new medicines for rare diseases^1^. The U.S. Food and Drug Administration (FDA) defines natural history as “the course a disease takes in the absence of intervention… from the disease’s onset until either the disease’s resolution or the individual’s death.”^1^ Such data may either arise from prospective observational research studies, or may be routinely collected as part of healthcare delivery (often termed real-world data, or RWD^2^), and can be used to quantitatively characterize the trajectory of a disease phenotype under standard of care, or often, without any effective treatment at all. At a minimum, natural history data are critical to designing well-powered clinical trials. These data may, for instance, help to identify quantitative clinical endpoints, refine the inclusion/exclusion criteria to select patients likely to benefit, determine the duration of trial needed to observe a therapeutic effect, and estimate the number of patients needed for adequate statistical power under various assumptions. When certain criteria are met^3^, regulators sometimes even accept natural history data to serve as an external control arm (ECA), to which patients treated with an investigational drug may be compared. Regulatory considerations include the rarity and severity of the indication, as well as the adequacy of available natural history data.

Prion disease is a rare, uniformly fatal neurodegenerative disease for which the hopes for a first effective therapy have increased in recent years due to the validation of prion protein (PrP) lowering as a therapeutic hypothesis^4^ and the advancement of multiple drug candidates towards clinical trials^5^. Considering its rarity, prion disease is highly studied and is well-ascertained by national surveillance centers, but diagnostic delay can be long^6^ and clinical presentation heterogeneous^7^. Past randomized clinical trials in prion disease^8,9^ accepted patients even with very advanced disease^10^, and were designed using simple power calculations based on the expected clinical benefit from open-label studies, which ultimately proved not to replicate in a rigorous placebo-controlled design. More recently, the MRC Prion Disease Rating Scale (MRC-PDRS)^11–14^ has been used to quantify initial level of impairment and subsequent decline in an open label treatment setting^15^, offering hopes for more rational inclusion criteria and endpoints. Nonetheless, despite published survival models^16,17^ and simulations of prion disease trials^18^ and a Phase 1/2a trial of a PrP-lowering antisense oligonucleotide^19^, neither the specific trial designs contemplated by trial sponsors, nor the data that would support decisions around such designs, have been made public.

Many studies in prion disease have focused on other forms of natural history data, including the diverse phenotypic manifestations of prion disease^20,21^, the diagnostic and prognostic utility of biomarkers in individuals referred for diagnostic testing for prion disease^22–26^, and age-dependent disease penetrance^27,28^ as well as biomarker trajectories^29,30^ in individuals at risk for genetic prion disease. Here, our focus is on time-to-event data in the symptomatic course of prion disease. We reason that clinical trials will enroll patients only after some clinical milestone, such as presentation or diagnosis, and will follow them until some event defined as a primary endpoint, such as death, the onset of akinetic mutism, or some minimum cognitive test score. Characterizing the state of the published literature regarding the time intervals between these milestones can help on one hand to quantify the natural history of prion disease, and on the other hand, to point out deficiencies in available published data and identify specific needs for additional datasets to enable clinical trial modeling and design.

Accordingly, we aggregate data on the natural history of prion disease from a systematic literature search spanning 25 years of published clinical medicine papers. We extract group summary statistics as well as individual patient data into a public dataset, evaluate biases and deficiencies in the literature, and quantify the effects of covariates on time intervals. Our data provide a comprehensive characterization of what is published about prion disease natural history, while demonstrating that the existing literature provides inadequate data for trial design, pointing to a need for larger and richer datasets of individual patient data.

## Results

We designed a systematic search strategy (see Methods) for PubMed articles from 2000 - 2024 containing clinical data on at least 5 prion disease patients, including at least one time-to-milestone interval, such as the time from symptom onset to death. Of 660 articles meeting our search criteria, 245 after manual curation met our inclusion criteria (Figure 1). 118 (48%) of these papers provided any individual patient-level data (PLD), totaling 1,418 individual patients. 206 (84%) reported summary statistics on groups of patients (hereafter, aggregated data), totaling 682 cohorts, with 418 cohorts reporting median values. Both PLD and aggregated data exhibited publication bias towards genetic and acquired cases (Figure 1B-C). Incident cases are ∼85% sporadic, ∼15% genetic, and <1% acquired^31,32^, while both genetic and acquired etiologies were over-represented in literature reports.

**Figure 1.**
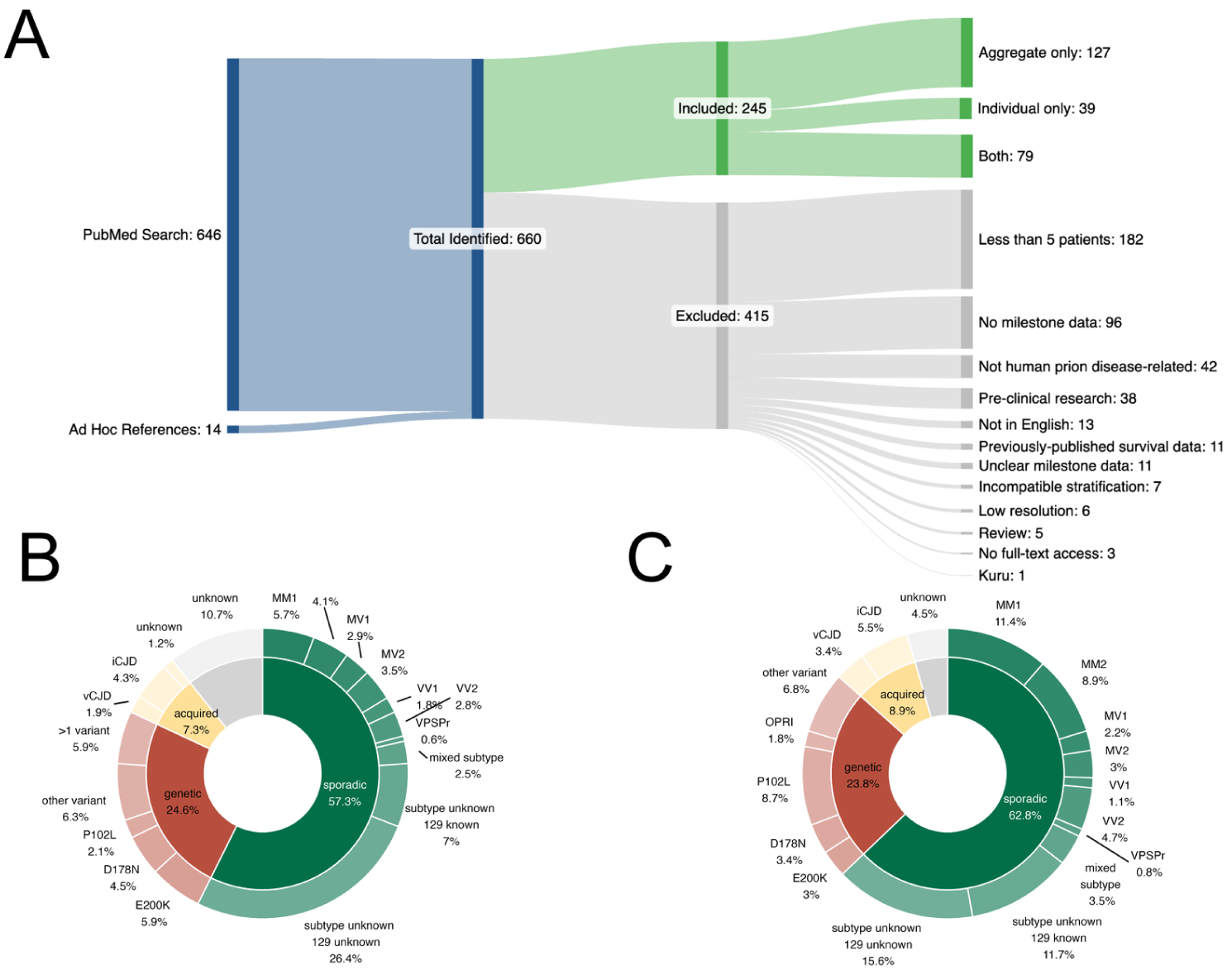
Search results and data extracted. **(A)** Sankey diagram of PubMed search (n=646) and ad hoc references (n=14) which identified 660 total papers. 245 papers were included, consisting of aggregate-only data (n=127), individual-level data (n=39), or both (n=79), resulting in a total dataset of 682 aggregate cohorts and 1,418 individual patients. **(B)** Etiological breakdown of aggregated cohorts are represented in the inner circle, and further subdivided into genetic variants and molecular subtypes in the outer ring. **(C)** Etiological classification of deduplicated patient-level data.

The various first presenting symptoms of prion disease^20^ can be followed by a months-long diagnostic odyssey^6^, resulting in a substantial interval between the time of symptom onset, retrospectively assessed through interview of family members, and the time of clinical presentation. Among aggregated data with a median time interval value reported, only 31/418 (7%) of cohorts had a time interval from a clinical presentation milestone (first evaluation or presentation, diagnosis, diagnostic test, sample collection, or study visit) to death (or akinetic mutism), with a weighted median of 46 (95% CI: 28-61) days (Table 1). Much more commonly, in 358/418 (86%) of cohorts, only the time interval from symptom onset to death was reported. The weighted median of time from symptom onset to death was 152 (95% CI: 152-152, N=378) days. This may suggest that typically 106 days, or 70% of the disease course, is spent in diagnostic odyssey. The aggregated cohorts exhibited substantial differences in survival time from symptom onset to death, depending upon the method by which the patients were ascertained: retrospective clinical cohorts (for instance, medical records searches or case series at a hospital) had the shortest survival, of 122 (95% CI: 122-137) days, while patients in prospective studies survived 70% longer, 207 (95% CI: 207-362) days. Cognitive tests were reported for only 41/378 (11%) of cohorts, which also trended towards longer-surviving patients, with survival of 183 (95% CI: 183-207) days versus 152 (95% CI: 149-152) days for those without cognitive tests.

**Table 1:** Biases and sources. Different variables affect the reported survival of aggregated cohorts, including method of ascertainment and/or the presence of a cognitive test. Weighted survival represents the weighted median of reported median onset to death intervals. N cohorts reflect the number of cohorts reporting median, while weighted time is a weighted median of reported medians. N individuals reflect the number of patients with study characteristics who also have a reported time interval. ^1^”Symptom” here refers to symptom onset. Reports of disease duration without a definition of the starting moment were presumed to be symptom onset. ^2^”Presentation” here includes first evaluation or presentation, diagnosis, diagnostic test, sample collection, or study visit. ^1,2^”Death” here refers to death and akinetic mutism as endpoints. ^3^Includes cohorts or individuals for which only clinical milestone data (such as those in Table 2) were reported. ^4^Includes only cohorts with a symptom to death time interval reported.

|  |  | aggregate |  |  |  | individual |  |  |  |
| --- | --- | --- | --- | --- | --- | --- | --- | --- | --- |
|  | study characteristics | time interval (days, weighted median) | 95% CI (days) | N cohorts | N PMIDs | time interval (days, median) | IQR (days) | N individuals | N PMIDs |
| survival interval reported | symptom – death<sup>1</sup> | 152 | 152-152 | 378 | 102 | 274 | 122-609 | 1287 | 109 |
|  | presentation – death<sup>2</sup> | 46 | 28-61 | 31 | 19 | 122 | 42-304 | 161 | 20 |
|  | other | 116 | 61-131 | 3 | 2 | 61 | 30-122 | 9 | 1 |
|  | none<sup>3</sup> | - | - | 26 | 10 | - | - | 82 | 12 |
|  | total unique | - | - | 418 | 122 | - | - | 1418 | 118 |
| ascertainment<sup>4</sup> | surveillance | 152 | 149-152 | 267 | 55 | 244 | 107-457 | 578 | 54 |
|  | retrospective | 122 | 122-137 | 64 | 27 | 271 | 122-883 | 469 | 37 |
|  | prospective | 207 | 207-362 | 31 | 13 | 362 | 163-723 | 86 | 10 |
|  | clinical trial | 524 | 472-524 | 2 | 1 | 222 | 117-502 | 51 | 3 |
|  | other, unknown, or multiple | 183 | 170-198 | 14 | 7 | 548 | 304-807 | 103 | 5 |
|  | total unique | - | - | 378 | 102 | - | - | 1287 | 109 |
| cognitive test<sup>4</sup> | no | 152 | 149-152 | 337 | 86 | 274 | 122-639 | 1207 | 104 |
|  | yes | 183 | 183-207 | 41 | 18 | 244 | 122-487 | 80 | 12 |
|  | total unique | - | - | 378 | 102 | - | - | 1287 | 109 |

Of the minority of aggregated cohorts for which a clinical presentation or diagnostic procedure was reported, the time intervals again varied considerably depending upon the nature of the milestone (Table 2). MRIs and lumbar punctures (LPs) were both the most common and occurred the earliest in the disease course, just 71 (95% CI: 65-91) and 83 (95% CI: 62-93) days after symptom onset respectively, while less common PET scans occurred 195 days after symptom onset. Similar trends were observed in the individual patient data, albeit with longer intervals, perhaps reflecting ascertainment bias in which patients had individual data reported. The different diagnostic tests and milestones differed not only in the time from symptom onset to test, but also in the time from symptom onset to death, suggesting they may have selected for slower or more rapid disease subtypes (Table 2).

**Table 2:** Survival and diagnostic delay by milestone. Weighted time represents the weighted median of reported median interval of onset to milestone. Weighted survival represents the weighted median of reported median onset to death intervals. N cohorts reflect the number of cohorts reporting a milestone in median. Where cohorts or individuals had >1 milestone reported, all milestones are included here, meaning those cohorts or individuals may count towards >1 row of this table. *LP includes both date of LP and date of CSF accessioning or analysis. ^1^Treatment indicates initiation of an investigational drug.

|  | procedure | aggregate |  |  |  |  |  | individual |  |  |  |  |  |
| --- | --- | --- | --- | --- | --- | --- | --- | --- | --- | --- | --- | --- | --- |
|  |  | Weighted median (onset to procedure, days) | 95% CI (days) | Weighted median (onset to death, days) | 95% CI (days) | *N* cohorts | *N* PMIDs | median time (onset to procedure, days) | IQR (days) | median time (onset to death, days) | IQR (days) | *N* individuals | *N* PMIDs |
| by procedure | MRI | 71 | 65-91 | 152 | 128-195 | 35 | 13 | 183 | 83 - 304 | 448 | 241-708 | 247 | 17 |
|  | LP* | 83 | 62-93 | 180 | 166-259 | 34 | 12 | 183 | 83 - 335 | 548 | 234-852 | 183 | 16 |
|  | EEG | 100 | 79-213 | 170 | 170-426 | 7 | 7 | 107 | 61 - 274 | 228 | 91-647 | 191 | 12 |
|  | treatment<sup>1</sup> | 125 | 95-152 | - | - | 4 | 2 | 183 | 107 - 320 | 557 | 178-1111 | 27 | 3 |
|  | PET/SP<br>ECT | 195 | 91-195 | 396 | 125-396 | 2 | 2 | 213 | 116 - 414 | 548 | 380-837 | 59 | 4 |
|  | neurological or<br>cognitive test | 304 | 213-731 | - | - | 3 | 1 | 122 | - | 304 | - | 1 | 1 |
|  | none | - | - | - | - | 346 | 99 | - | - | - | - | 972 | 92 |
|  | *total*<br>*unique* | - | - | - | - | 418 | 122 | - | - | - | - | 1418 | 118 |
| by<br>clinical<br>milestone | presenta<br>tion | 61 | 56-153 | 166 | 148-166 | 9 | 5 | 122 | 54 - 213 | 304 | 118-434 | 142 | 15 |
|  | diagnosi<br>s | 113 | 83-113 | 131 | 125-131 | 22 | 11 | 61 | 35 - 146 | 244 | 95 - 320 | 31 | 5 |
|  | study<br>visit | 131 | 122-183 | 390 | 390-441 | 17 | 5 | - | - | - | - | - | - |
|  | none | - | - | - | - | 374 | 104 | - | - | - | - | 1254 | 107 |
|  | *total*<br>*unique* | - | - | - | - | 418 | 122 | - | - | - | - | 1418 | 118 |

Survival curves were estimated from time of symptom onset to death constructed from PLD, stratified by histologic subtype^33^. Results replicated previously reported differences in survival^34^ among these subtypes (Figure 2A). Median survival was longer when the endpoint was considered to be death only (Figure 2A), versus an endpoint of either death or akinetic mutism (Figure 2B). Among N=55 sCJD MM1 individuals with both of these endpoints reported, the median time was 91 days to akinetic mutism and 335 to death, a difference of 244 days (p=1.5E-14, log-rank test). Thus, among the total of N=150 MM1 individuals in the dataset, the median time to endpoint was 91 days when akinetic mutism or death were considered, while it was 152 days when death only was considered, a difference of 61 days (p=3.7E-4, log-rank test). Similarly for MM2, the N=22 individuals with both endpoints reported survived 563 days to akinetic mutism and 852 days to death (p=0.042, log-rank test), resulting in the overall distribution for N=118 MM2 individuals shifting from a median of 700 to 731 days (p=0.53, log-rank test) . This quantifies the degree to which life-extending measures, which are particularly common in Japan^35^, where nearly all patients have a 129MM genotype^36^, inflate survival time in prion disease.

**Figure 2.**
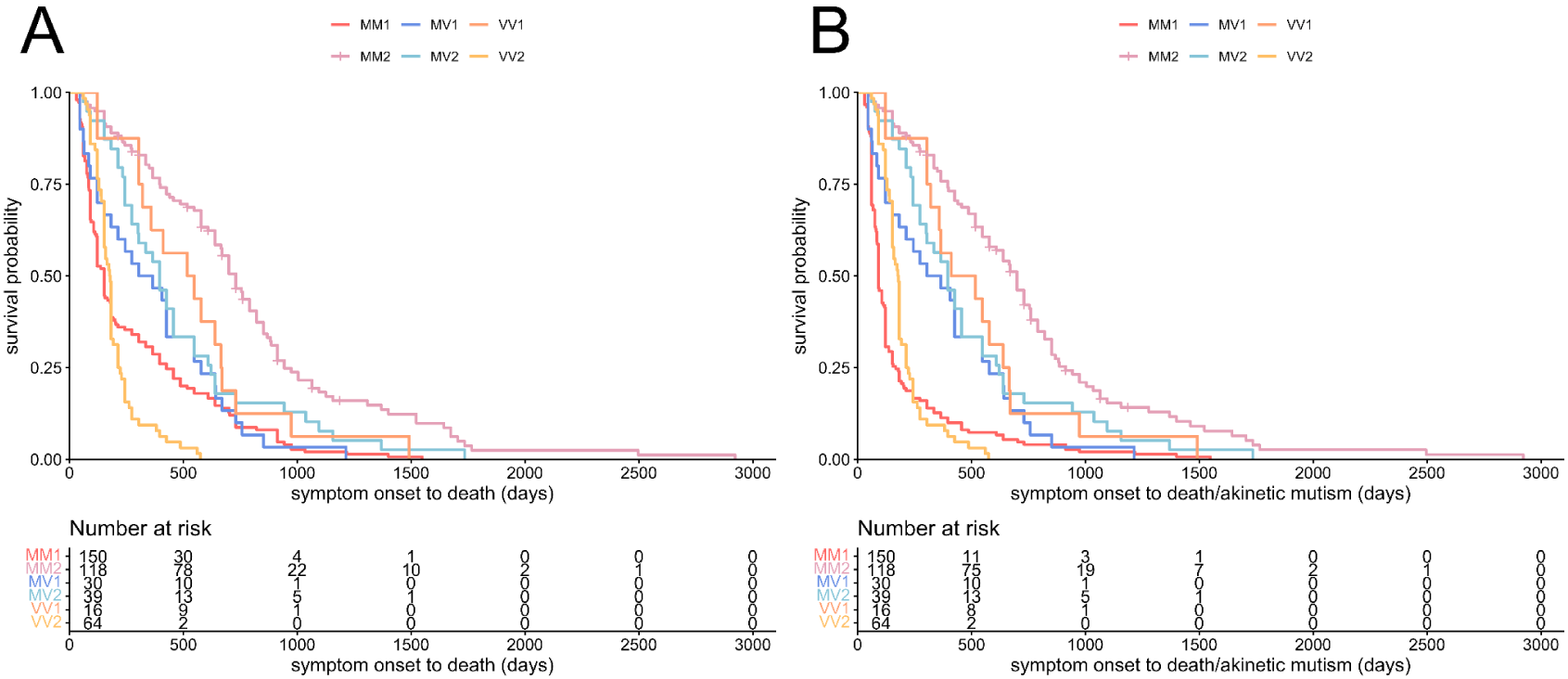
Survival of individual sporadic CJD patients by histopathologic subtype. Kaplan-Meier curve of individual patients with sporadic CJD (N=417). Survival curves shift to the left when the endpoint is adjusted to prioritize time from onset to akinetic mutism **(B)** over onset to death **(A)** when both are reported.

In univariate Cox proportional hazards analyses of individual patient data, we replicated the reported^16^ shorter survival from symptom onset to death or akinetic mutism in males (hazard ratio HR = 1.16, P = 0.024) and in older individuals (HR = 1.04 per year, P = 4.1E-35) (Table 3). Even after controlling for these demographic variables in multivariate Cox models, we observed highly significant differences in survival between disease subtypes for both genetic and sporadic disease (Table 3), replicating reported^27,34^ differences. These subtype differences were likewise reflected in the distribution of reported median survival times from symptom onset to death or akinetic mutism for aggregated cohorts for both sporadic (Figure 3A) and genetic (Figure 3B) prion disease. At the same time, the spread of the aggregated cohort medians — a 3.2-fold difference between highest (776 days) and lowest (244 days) median survival time for MM2, for example — may reflect differing ascertainment methods (Table 1) selecting for patients with dramatically different survival times. A mixed-effect meta-regression on median survival values of aggregate cohorts by definite or probable histological subtype indicated a very large amount of unexplained heterogeneity (I^2^ = 95.5%)^37^.

**Figure 3.**
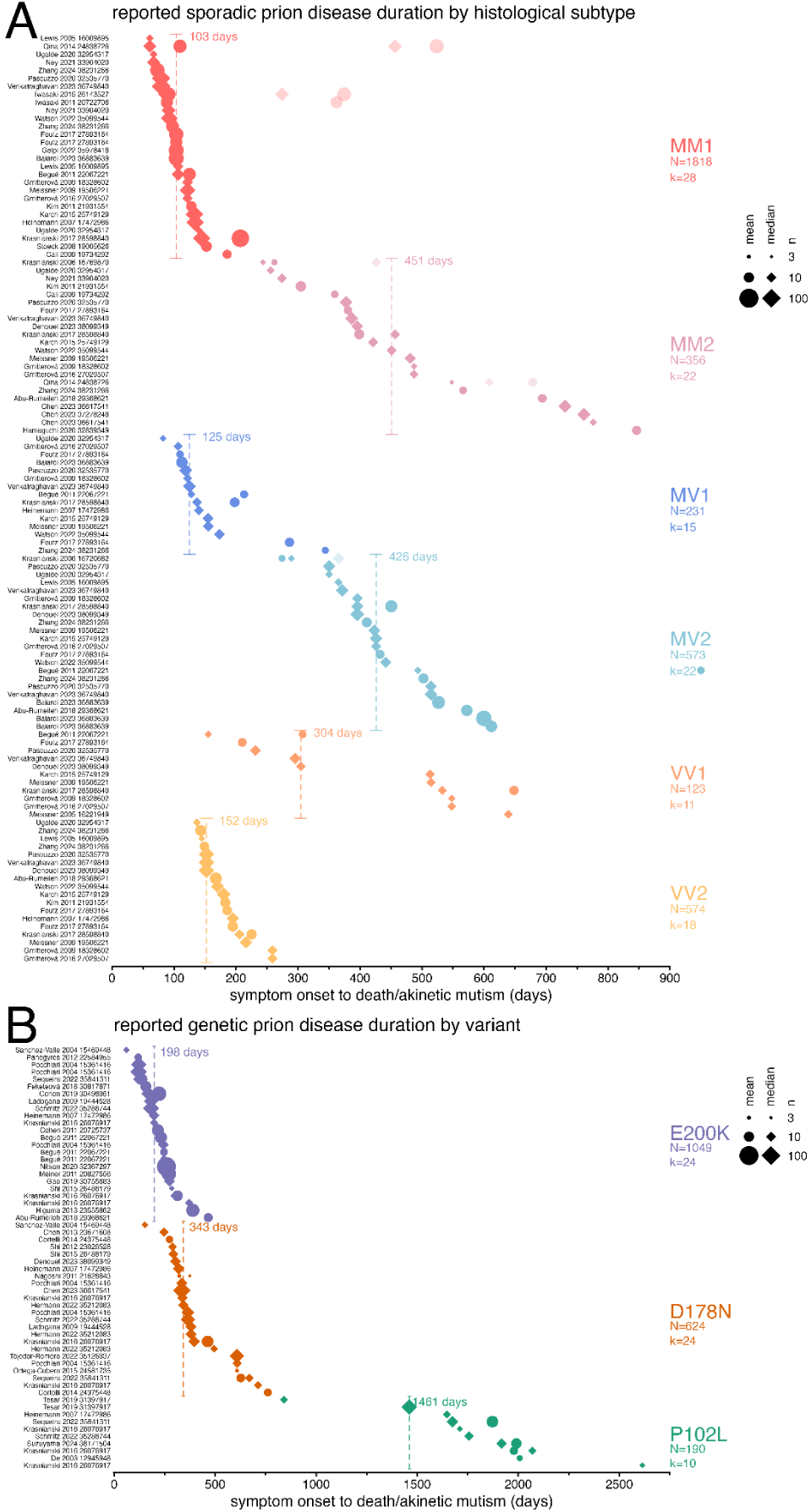
Forest plot of reported prion disease survival by prion disease subtype. Survival time from symptom onset to death or akinetic mutism for aggregated cohorts by sporadic CJD histological subtype **(A)** or genetic prion disease variant **(B)**. Data points represent medians or, where medians were unavailable, means. The symbol size is proportional to the cohort size. An akinetic mutism endpoint (n≥2 patients per cohort) is prioritized where available, and if akinetic mutism was reported, then data for a death endpoint (n≥3 patients per cohort) are plotted with higher transparency. N indicates total individuals and k indicates the number of unique cohorts per subtype. Dashed vertical lines indicate the weighted median survival for each subtype.

**Table 3.** Cox proportional hazards models fit on individual patient data. MM1 reference group for subtype sporadic analysis. D178N reference group for genetic analyses. Hazard ratios for age of onset represent a per year change.

| dataset | Cox model type | term | N events/total | hazard ratio (95% CI) | P value |
| --- | --- | --- | --- | --- | --- |
| sporadic and genetic | univariable | age of onset (per yr) | 691/738 | 1.04 (1.03-1.05) | 4.10E-35 |
|  | univariable | sex: male | 978/1024 | 1.16 (1.02-1.31) | 2.40E-02 |
|  | univariable | sCJD | 1070/1119 | 2.97 (2.54-3.47) | 7.60E-43 |
|  | multivariable | sCJD | 656/700 | 2.27 (1.87-2.76) | 1.70E-16 |
|  |  | age of onset | 656/700 | 1.03 (1.03-1.04) | 3.80E-19 |
|  |  | sex: male | 656/700 | 1.18 (1.02-1.38) | 3.10E-02 |
| sporadic | univariable | age of onset (per yr) | 459/483 | 1.03 (1.02-1.04) | 1.80E-12 |
|  | univariable | sex: male | 719/742 | 1.10 (0.95-1.27) | 2.20E-01 |
|  | multivariable | MM2 | 296/311 | 0.08 (0.06-0.11) | 1.50E-43 |
|  |  | MV1 | 296/311 | 0.14 (0.08-0.23) | 2.10E-14 |
|  |  | MV2 | 296/311 | 0.15 (0.09-0.24) | 5.40E-14 |
|  |  | VV1 | 296/311 | 0.16 (0.08-0.33) | 5.80E-07 |
|  |  | VV2 | 296/311 | 0.58 (0.40-0.83) | 2.70E-03 |
|  |  | age of onset (per yr) | 296/311 | 1.03 (1.01-1.04) | 3.50E-05 |
|  |  | sex: male | 296/311 | 1.49 (1.17-1.89) | 1.10E-03 |
| **genetic** | univariable | E200K | 282/306 | 1.49 (0.97-2.29) | 6.70E-02 |
|  | univariable | other variant | 282/306 | 0.54 (0.37-0.80) | 1.70E-03 |
|  | univariable | P102L | 282/306 | 0.33 (0.23-0.48) | 6.50E-09 |
|  | univariable | age of onset (per yr) | 232/255 | 1.03 (1.02-1.05) | 1.10E-10 |
|  | univariable | sex: male | 259/282 | 1.30 (1.01-1.66) | 3.80E-02 |
|  | multivariable | E200K | 219/241 | 0.91 (0.55-1.50) | 7.00E-01 |
|  |  | other variant | 219/241 | 0.40 (0.25-0.62) | 4.60E-05 |
|  |  | P102L | 219/241 | 0.22 (0.14-0.34) | 8.40E-12 |
|  |  | age of onset (per yr) | 219/241 | 1.04 (1.03-1.06) | 5.60E-13 |
|  |  | sex: male | 219/241 | 0.96 (0.73-1.27) | 7.80E-01 |

## Discussion

Prion disease is widely studied. We were able to identify 245 medical articles that reported time-to-event or cognitive test data on prion disease patients. Despite this, our literature search illustrates that the existing literature does not provide an adequate characterization of the natural history of the symptomatic progression of prion disease sufficient to guide the design clinical trials, let alone serve as an external control arm. Several limitations contribute to this problem.

The most valuable types of data for designing clinical trials are unfortunately the most uncommon. The ideal dataset would consist of longitudinal PLD quantifying disease severity and progression between the time of diagnosis and death. Instead, we find that data in the literature are overwhelmingly cross-sectional rather than longitudinal, aggregated rather than individual, use symptom onset rather than diagnosis as a start time, and lack any metrics of disease severity.

The literature focuses disproportionately on the total survival time from first symptom to death. 90% of cohorts reported time intervals beginning with symptom onset, compared to 7% beginning with a clinical presentation milestone. This symptom-to-death time interval may be relevant for trials of pre-symptomatic individuals at genetic risk of prion disease, but is of limited utility for clinical trial design in already-symptomatic individuals, because patients are not available to enroll in trials from the first moment they experience a symptom. Indeed, the data we extracted suggest that ∼70% of the disease course has historically been spent in diagnostic odyssey (106 out of 152 days). Moreover, both the start and end time of this interval are fuzzy. Symptom onset is assessed by retrospective interview with family members, who may not always remember clearly or agree with one another what happened. The time of death is determined not only by underlying disease biology but also by family decisions around end-of-life care, as illustrated by the 244 day difference between median time to death and time to akinetic mutism for MM1 patients with both endpoints reported in this dataset. More useful for clinical trials would be the time interval from diagnosis to either death or life-extending measures. Unfortunately, in past randomized clinical trials in prion disease, many patients were already on life-extending measures at the time of enrollment^10^, suggesting a need to set inclusion criteria selecting for patients with some quality of life remaining, and earlier disease endpoints that may reflect preservation of this quality of life.

Overall survival is not the only data required for a well-designed clinical trial. An estimate of disease course is essential to developing drugs with a patient endpoint other than just death. Unfortunately, the literature lacks longitudinal data of disease burden. Only 41 cohorts and 80 individual patients with survival outcomes reported also had any cognitive testing performed, and of whom only 12 individuals had >1 timepoint of cognitive test results reported. Thus, while some limited data are available on the time from diagnosis to death, it is impossible to quantify how impaired those patients were at the time of diagnosis, or how long prior to death they might have progressed beyond some minimum cognitive or functional score.

We found evidence of substantial publication bias and ascertainment bias. Rare prion disease subtypes, particularly acquired prion disease, are over-represented in literature reports, and the survival time of patients varies widely depending upon how the cohort of patients was ascertained. Surveillance and retrospective cohorts, which are likely to catch a wide net of patients, reported shorter survival times than prospective cohorts, in which the only patients who survive long enough to enroll are those who are either lucky to be diagnosed very early, or have slower-progressing subtypes. This likely contributed to as much as a 3.2-fold difference in survival between reported cohorts, even after controlling for histologic subtype. Clinical trial participants are likely to more closely mirror prospective cohorts. Comparison of trial participants to surveillance cohorts, where survival may be 71% shorter, may be one reason why some open label studies of doxycycline and quinacrine in prion disease were interpreted to suggest a clinical benefit that was never borne out by randomized trials^8,9^.

The literature predominantly reports aggregated data. We extracted PLD for 1,418 deduplicated individual patients, however, these were even more heavily focused on symptom-to-death intervals, with time from diagnosis to death reported for just 1.9% (N=27). Given that the timing of diagnosis, disease stage or severity at time of diagnosis, and rate of progression after diagnosis are not reported for most individuals in our PLD dataset, these data are likely not adequate for modeling out clinical trial scenarios.

Putting together the above constraints, it is clear that clinical trial sponsors would be hard-pressed to design a clinical trial using data available in the literature today. A reasonable sponsor might seek to enroll patients who remain above some minimum MRC-PDRS cutoff at time of diagnosis, who are expected to survive at least the ∼1 month required for PrP-lowering drugs to take full effect^38^, and might wish to know what percent of such patients will reach an endpoint of death or life-extending measures or will remain above some minimum MRC-PDRS value within a specified number of weeks. Here we have extracted a comprehensive dataset of published PLD and aggregated data from the literature, but these data are not sufficient to model out the above assumptions.

Our study has limitations. Here, we only collected data present in the main text or supplementary tables of published papers. We did not systematically contact the corresponding authors of all such papers to request raw underlying data. We also focused on papers reporting at least 5 patients and did not extract individual case reports. Heterogeneity in data reporting practices — for example, some aggregated cohorts had means while others had medians — further limited the comparisons we could make. Inherent limitations of our systematic literature review also include: lack of detail on ascertainment method or inclusion criteria for many papers; inability to de-duplicate patients in aggregated data and imperfect ability to de-duplicate individual data; variable missingness of information on genotype, disease subtype, biomarker values, and demographics.

Despite these limitations, the dataset that we release here comprises, to our knowledge, the largest public dataset available characterizing the natural history of the symptomatic course of prion disease.

We highlight several priorities for the prion field. First, public deposition of appropriately de-identified PLD should become the norm and expectation. PLD are essential for designing and modeling clinical trials, controlled-access databases are often unavailable to industrial sponsors, and survey data shows that research participants are generally supportive of sharing their data^39,40^. Public PLD deposition should be anticipated in consent forms, required for publication of journal articles, and made a condition of funding by patient organizations and national funding agencies. Second, authors reporting clinical data should be encouraged to report clinically relevant parameters, such as time interval from presentation or diagnosis to death or life-extending measures, rather than only time from symptom onset to death. Third, clinicians and investigators should collect, and report, cognitive or functional scores including MRC-PDRS at first assessment.

All clinical data ultimately come from patients, and our duty to do right by patients demands that we make high quality data available to guide the rational design of well-powered trials that will ultimately make prion disease a treatable condition.

## Methods

### Study design

Our goal was to characterize the existing literature regarding the natural history of prion disease from symptom onset to death, in order to provide a quantitative basis for planning and design of future clinical trials. We sought to include, where available, milestones such as referral, clinical diagnosis, cohort enrollment, diagnostic tests, and information on diagnostic test results as well as autopsy results and prion disease subtypes. We set out to accomplish this through a systematic literature search, extraction of quantitative data from published papers, and analysis and public release of these quantitative data. We followed the PRISMA guidelines for systematic reviews^41^. The review was not pre-registered.

### Search strategy and selection criteria

We searched PubMed on December 24, 2024, and selected papers published since January 1, 2000. The search string was “(”prion disease”[Title/Abstract] or “prion diseases”[Title/Abstract] or “CJD”[Title/Abstract] or “familial fatal insomnia”[Title/Abstract] or “Creutzfeldt-Jakob disease” [Title/Abstract] or “Transmissible spongiform encephalopathy”[Title/Abstract] or “Gerstmann-Straussler-Scheinker”[Title/Abstract] or “prionopathy”[Title/Abstract] or “VPSPr” [Title/Abstract]) AND (”natural history” or “duration” or “survival” or “progression” or “surveillance”) AND (“patients”[Title/Abstract] or “cohort”[Title/Abstract] or “patient”[Title/Abstract] or “cases”[Title/Abstract] or “surveillance”[Title/Abstract]) NOT (Review[Publication Type]) NOT “cancer” NOT “oncology” NOT (“mouse”[Title]) NOT (“mice”[Title])”. A review protocol was not prepared. Ad hoc references were added where reference lists in the identified reports suggested additional relevant titles. Titles and abstracts were then screened by one reviewer (LMHX) according to pre-specified inclusion/exclusion criteria (Figure 1A). Eligible papers were required to have full text available in English, quantitatively report original human data, and to report a time-to-milestone for N≥5 prion disease patients. Of 660 titles reviewed, we excluded studies reporting less than 5 patients (N=182), lacked time-to-milestone data (N=96), were not related to human prion disease (N=42), were pre-clinical (N=38), reported previously-published data (N=11), had unclear survival or milestone data (e.g. summary statistics visualized but not presented in tabular form and so exact numeric values unavailable; N=11), were not in English (N=13), had an incompatible survival stratification (e.g. prion disease grouped with other diagnoses; N=7), had low resolution of outcome times (e.g. years instead of months; N=6), were review articles (N=5), or for which we could not access full text (N=3). Because none of the other included titles reported patients with kuru, we decided to exclude a title reporting exclusively on kuru (N=1).

### Data extraction

We sought quantitative data on time-to-milestone, survival, disease etiology and histologic subtype or *PRNP* variant, codon 129 genotype, cognitive test scores, ascertainment, sample size, sex, autopsy results, biomarker values (real-time quaking-induced conversion or RT-QuIC, total tau or T-tau) and age. Available data were manually extracted by 1 reviewer (LMHX) into two spreadsheets, one for aggregate cohorts (grouped patient data with n≥2 patients) and one for individual patient data on Google Sheets. In the case where an n=2 aggregate cohort reported time-to-milestone as a range, the patients were also captured in the individual dataset. Where disease duration was reported without an explicit statement of the start time of the interval, the start was assumed to be symptom onset. Where the ascertainment approach was not stated, we made assumptions based on the authors’ affiliations, for instance, publications from prion surveillance centers were assumed to be surveillance data. Diagnostic milestones were grouped as follows: diagnosis (including collection or accessioning of diagnostic sample, diagnostic testing, or referral to surveillance centers), presentation (including admission, referral to specific clinic or neurologist), and study visit. Missing quantitative values were left blank. Individuals were considered to have a cognitive test score only if the scores were reported; slopes (e.g. MRC-PDRS points per day) reported without the underlying scores did not count. Individual patients or cohorts found to contain errors were removed (examples: diagnostic test date prior to disease onset; genetic variant reported with wrong reference amino acid). All time intervals were harmonized into days; note that conversion of intervals reported in months resulted in a preponderance of time intervals of 30, 61, 91, 122, 152, or 183 days. Patient-level total tau levels were extracted when reported exactly and not when reported as above or below a threshold. Risk of bias in the studies was evaluated through statistical analyses reported within the Results, pertaining to the disease subtypes, ascertainment approaches, and times to event. Analyses indicated that bias is likely pervasive, and no studies were excluded based on risk of bias.

### Use of large language models (LLMs)

OpenAI Codex (5.5) was used to generate some of the source code for this study. LLM-generated code was reviewed, tested, and validated by LMHX. Claude Code (Opus 4.8) was further used to audit source code. We also evaluated the possibility of automation by using LLMs to distill data from papers, but found that hallucinations were common and that cohort metadata not embedded within tables was not readily extracted, therefore, all data extraction was performed manually (see “Data extraction” above).

### Statistical analysis and data availability

Analyses utilized custom scripts in R 4.4.3. The sankey diagram in Figure 1 was produced using sankeymatic.com; source data are also available in the git repository. Kaplan-Meier survival analyses were performed with the R packages survival 3.8.3 and survminer 0.5.0, and Cox proportional hazards analyses were conducted with survival 3.8.3. Meta-regression was conducted with the R package metamedian^42^ 1.2.1 using a quantile matching estimation method to estimate the pooled median. Analyses were conducted using a random-effects model, and heterogeneity statistics were estimated via Restricted Maximum Likelihood. Weighted medians and confidence intervals of milestone time intervals were also calculated with metamedian using sample size as the weight. Statistics in tables 1 and 2 are descriptive. We used reported time-to-milestone median values and excluded mean values with the assumption that reported durations were not normally distributed. Analyses of aggregate cohorts were limited to cohorts with n≥3 patients.

### Individual patient data deduplication

Because the same individual patients may be reported in more than 1 study, potential duplicate individual records were identified using a custom R script. Entries sharing identical values for survival duration, sex, age of onset, region, and subtype were flagged; only one record per duplicated set was retained for subsequent analyses. Aggregate cohort data are also certain to contain duplicates, but in this case no deduplication is possible based on reported data.

### Glycotype Conversion

Reported PrP glycotypes were standardized according to the Parchi classification system.^24^ Glycotypes reported under the London classification system^25^ were mapped to the Parchi system as such: London types 1 and 2 to Parchi type 1, and London type 3 to Parchi type 2.

### Source code and data availability

All data extracted from the literature as well as source data and summary statistics for figures and tables herein are available in the supplemental data. The curated dataset and source code sufficient to reproduce all analyses herein are also available in a public git repository: http://github.com/vallabhminikel/nat_hist_literature_data

## Data Availability

Analyses utilized custom scripts in R 4.4.3. The sankey diagram in Figure 1 was produced using sankeymatic.com; source data are also available in the git repository. Kaplan-Meier survival analyses were performed with the R packages survival 3.8.3 and survminer 0.5.0, and Cox proportional hazards analyses were conducted with survival 3.8.3. Meta-regression was conducted with the R package metamedian42 1.2.1 using a quantile matching estimation method to estimate the pooled median. Analyses were conducted using a random-effects model, and heterogeneity statistics were estimated via Restricted Maximum Likelihood. Weighted medians and confidence intervals of milestone time intervals were also calculated with metamedian using sample size as the weight. Statistics in tables 1 and 2 are descriptive. We used reported time-to-milestone median values and excluded mean values with the assumption that reported durations were not normally distributed. Analyses of aggregate cohorts were limited to cohorts with n≥3 patients.

## Acknowledgements

SMV acknowledges speaking fees from Abbvie, Biogen, Eli Lilly, Illumina, Ultragenyx, and Voyager; consulting fees from Alnylam, Invitae, and Regeneron; research support from Cenos, Eli Lilly, Gate Bio, Ionis, and Sangamo Therapeutics. EVM has received speaking fees from Abbvie, Eli Lilly, Novartis, Vertex, and Voyager; consulting fees from Alnylam, Arrowhead, Deerfield, and Regeneron; and research support from Cenos, Eli Lilly, Gate Bio, Ionis, and Sangamo Therapeutics. The authors have no additional financial interests.

## Funding

This study was supported by the National Institutes of Health (R01 NS132022 to EVM) and by donations to the Prions@Broad fund at the Broad Institute.

## Notes

### Author Declarations

We extracted data from medical publications between 2000 and 2024 reporting on n≥5 prion disease patients. Information about demographics, ascertainment, and clinical milestones, such as time from onset to death, akinetic mutism, diagnostic testing and outcomes, were extracted for aggregate cohorts of ≥2 patients and individual patient-level data (PLD). We computed summary statistics of cohorts and PLD, visualized survival through forest plots and Kaplan-Meier curves, analyzed covariates regarding survival, and identified biases and heterogeneity within the literature.

